# Diagnosing and remediating harmful data shifts for the responsible deployment of clinical AI models

**DOI:** 10.1101/2023.03.26.23286718

**Authors:** Vallijah Subasri, Amrit Krishnan, Azra Dhalla, Deval Pandya, David Malkin, Fahad Razak, Amol A. Verma, Anna Goldenberg, Elham Dolatabadi

**Affiliations:** Genetics and Genome Biology Program, The Hospital for Sick Children, Toronto, ON, Canada; Department of Medical Biophysics, University of Toronto, Toronto, ON, Canada; Vector Institute for Artificial Intelligence, Toronto, ON, Canada; St Michael’s Hospital, Unity Health Toronto, Toronto, ON, Canada; Institute of Health Policy, Management and Evaluation (IHPME), University of Toronto, Toronto, ON, Canada; Department of Medicine, University of Toronto, Toronto, ON, Canada; Division of Hematology/Oncology, The Hospital for Sick Children, Toronto, ON, Canada; Department of Pediatrics, University of Toronto, Toronto, ON, Canada; Department of Computer Science, University of Toronto, Toronto, ON, Canada

## Abstract

Harmful data shifts occur when the distribution of data used to train a clinical AI system differs significantly from the distribution of data encountered during deployment, leading to erroneous predictions and potential harm to patients. We evaluated the impact of data shifts on an early warning system for in-hospital mortality that uses electronic health record data from patients admitted to a general internal medicine service, across 7 large hospitals in Toronto, Canada. We found model performance to differ across subgroups of clinical diagnoses, sex and age. To explore the robustness of the model, we evaluated potentially harmful data shifts across demographics, hospital types, seasons, time of hospital admission, and whether the patient was admitted from an acute care institution or nursing home, without relying on model performance. Interestingly, many of these harmful data shifts were unidirectional. We found models trained on community hospitals experience harmful data shifts when evaluated on academic hospitals, whereas models trained on academic hospitals transfer well to the community hospitals. To improve model performance across hospital sites we employed transfer learning, a strategy that stores knowledge gained from learning one domain and applies it to a different but related domain. We found hospital type-specific models that leverage transfer learning, perform better than models that use all available hospitals. Furthermore, we monitored data shifts over time and identified model deterioration during the COVID-19 pandemic. Typically, machine learning models remain locked after deployment, however, this can lead to model deterioration due to harmful data shifts that occur over time. We used continual learning, the process of learning from a continual stream of data in a sequential manner, to mitigate data shifts over time and improve model performance. Overall, our study is a crucial step towards the deployment of clinical AI models, by providing strategies and workflows to ensure the safety and efficacy of these models in real-world settings.

## Introduction

AI systems have leveraged clinical data to predict mortality ^1–5^, length of stay (LOS) ^6^, sepsis ^7–9^ and the occurrence of specific disease diagnoses^10^. As a growing number of AI systems are sought to be deployed in clinical settings, a defining challenge for AI in healthcare is how to responsibly deploy models that have been developed ^11,12^. Building robust clinical machine learning (ML) models has proven to be difficult ^13^, in part attributed to *data shifts (*or *data drift)*–changes in the data distribution over time and/or space that leads to spurious predictions^14^. This can occur due to changes in the *features* of the input data or due to changes in the *labels,* which represent the outcome the model is predicting. Data shifts are harmful when they result in *model drift–*a significant decrease in the model’s predictive power due to changes in the real world environment. A key barrier to the safe deployment of clinical AI systems is attributed to system malfunction due to harmful data shifts ^15^. Data shifts occur when the underlying distribution of the data used to build a predictive model differs from the distribution of the data encountered during deployment. In healthcare, these shifts can exist along the axes of institutional differences (e.g., staffing, instruments and data-collection workflows), epidemiological changes (e.g. diseases, catastrophic events)^16^, temporal shifts (e.g. policy changes, changes in clinician or patient behaviours over time)^17^ and differences in patient demographics (e.g. race, sex, age, socioeconomic background, and types of presenting illnesses and comorbidities)^18–20^. When the difference between the training and test data distribution is sufficient to deteriorate the model’s performance, clinical decision-making may be impaired. As a result, it is imperative to identify these potentially harmful shifts *a priori*, to inform clinical end-users and prevent harm to patients.

Rigorous evaluations across time, hospital sites, and patient characteristics are critical for identifying model degradation and ensuring equitable and quality patient care. The impact of distributional shifts on model performance ^21^ has been explored for the prediction of sepsis ^22^, mortality ^19,23^, ER admissions ^16^, LOS ^19^ and *Clostridioides difficile* infections ^17^. Model deterioration has previously been associated with transitions in EHRs systems over time ^13^ and across patient demographics in chest X-rays ^24^, skin lesions^25^ and sepsis prediction ^26^. However, in many clinical prediction problems, the lead time to acquire labels is lengthy, and the process is resource-intensive. Labels like death or sepsis are rare; this causes a delay in the ability to detect a statistically significant change in model performance, at which point model deterioration may have already occurred, and it may be too late to take steps for remediation. This suggests retraining based on recognizing deterioration in model performance is impractical, and emphasizes the importance of detecting harmful data shifts in a label-agnostic manner ^27–29^. Furthermore, it is necessary to design effective strategies for model updating that proactively minimize model degradation in the presence of data shifts. Failure to correct for harmful data shifts can lead to the perpetuation of algorithmic biases, missing critical diagnoses and unnecessary clinical interventions that can be detrimental to patient outcomes and burden the healthcare system^11,12^.

In this study, we developed an evaluation and monitoring pipeline to prepare clinical AI systems for deployment.^30^ We used our pipeline to monitor for harmful data shifts in a label-agnostic manner using an early warning system (EWS) for all-cause in-hospital mortality. In doing so, we proactively identified harmful data shifts across various real-life scenarios, including institutional differences, time of hospital admission, whether a patient was admitted from an acute care institution or nursing home and the COVID-19 pandemic. Interestingly, we found many of these harmful data shifts were unidirectional, suggesting that there exists unique patient populations in certain clinical contexts that are not captured in others. In the presence of harmful data shifts across institutions, we leveraged transfer learning to identify strategies for improving model performance ^31–33^. Lastly, we conducted a prospective evaluation, whereby we monitored for temporal data shifts and used continual learning to proactively update clinical AI models under harmful data shifts.

## Results

### All-cause in-hospital mortality early warning system (EWS)

We developed a dynamic EWS to predict the risk of in-hospital mortality within the next two weeks, every 24 hours, using EHR data consisting of lab results, transfusions, imaging reports and administrative features (**Supplementary Table 1-2; Supplementary Figure 1**) from 109,802 patient encounters admitted to general internal medicine (GIM) inpatient units at seven large hospitals in the greater Toronto area (GTA, in Canada). Given the varying distribution of diagnoses and demographics across hospitals (**Figure 1A-C**), we assessed the fairness of our model by evaluating the area under the receiver operating characteristic (AUROC) and area under the precision-recall curve (AUPRC) for subgroups of diagnoses, sex and age in our test data^34^. We defined diagnostic subgroups using the ICD-10 diagnosis chapters^35^–groupings of ICD-10 diagnosis codes assigned to patients during admission based on affected body systems and health conditions. We found that the model performed particularly well on certain diagnoses, including diseases of the circulatory system (I00-I99), respiratory system (J00-J99), COVID-19 (U07-U08) and certain other infectious and parasitic diseases (A00-B99). However, it had a much lower AUROC on individuals with benign or malignant neoplasms (C00-D49) and factors influencing health status and contact with health services (Z00_Z99; **Figure 1D**). These primarily consisted of patients receiving palliative care (n_Z515_=2042; **Supplementary Figure 2**), including patients with cancer, heart failure, chronic obstructive pulmonary disease (COPD), dementia, and Parkinson disease. This is in accordance with what we know about palliative care as encompassing complex diseases with evolving needs, caused by a combination of genetic, environmental and lifestyle factors, which may make it more difficult to accurately predict using EHR data^36^. We also found AUROC increased and AUPRC decreased across groups with decreasing age, this may be in part driven by the lower mortality rates in the younger age groups. Alternatively, performance was fairly consistent across sex (**Figure 1D**). We also compared the performance of our model, which included no prior information of patient history, to models that included comorbidities and ICD-10 diagnosis codes as features (**Supplementary Table 1**). In doing so, we found that including ICD-10 diagnosis codes as features in our model slightly improved overall performance (**Figure 1E**), but significantly increased the performance gap between many diagnostic subgroups (**Supplementary Figure 3**).

**Figure 1.**
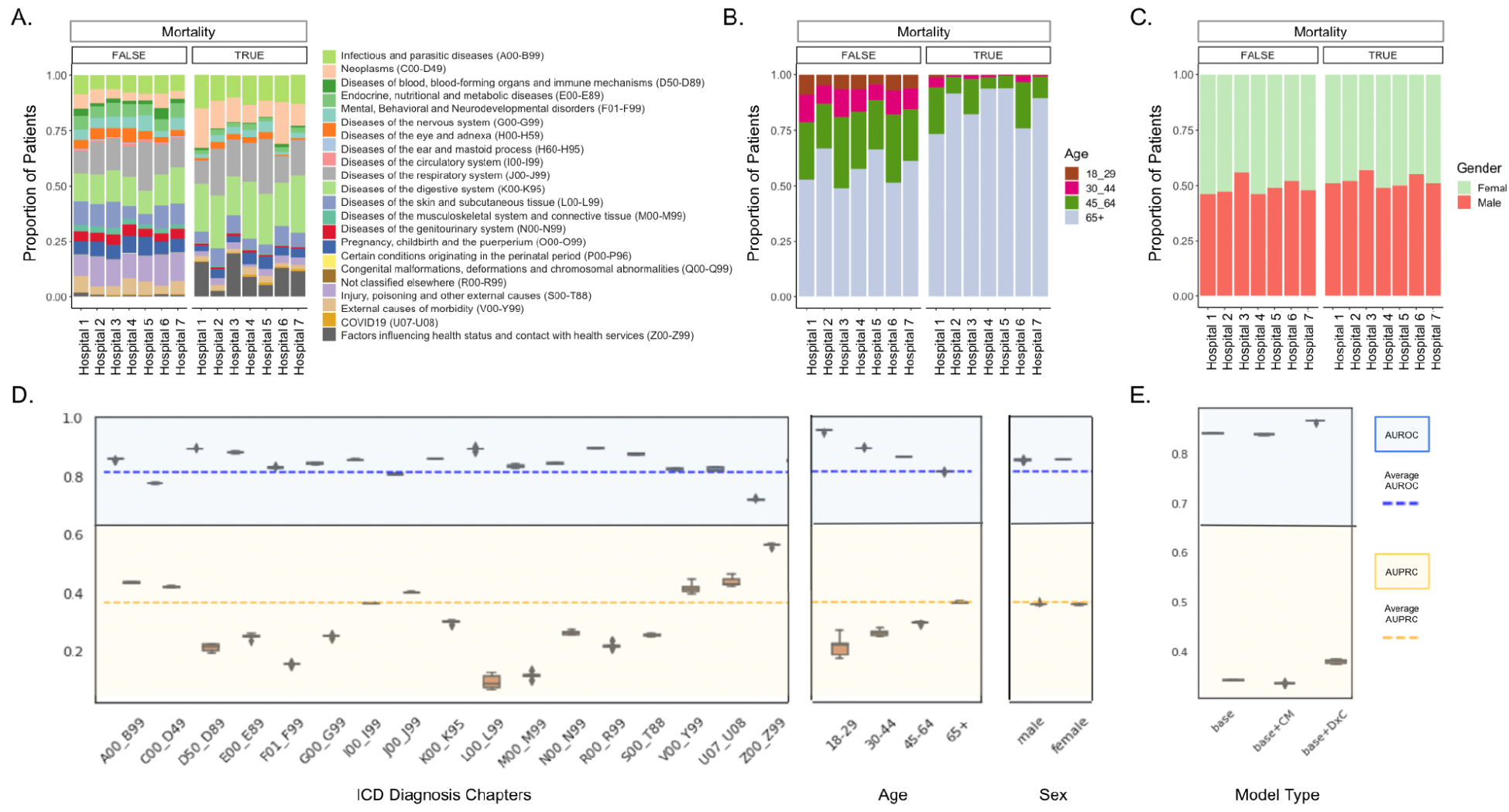
Patient characteristics and model fairness across subgroups. **(A)** Distribution of ICD-10 diagnosis codes across hospitals by mortality status (true/false). **(B**) Distribution of age groups across hospitals by mortality status (true/false). **(C)** Distribution of sex across hospitals by mortality status (true/false). **(D)** AUROC and AUPRC of cross-site EWS across ICD-10 diagnosis codes, age and sex. **(E)** Overall AUROC and AUPRC of the model without prior information (base), with comorbidities (base+CM) and with diagnosis codes (base+DxC) as features.

**Table 1.**
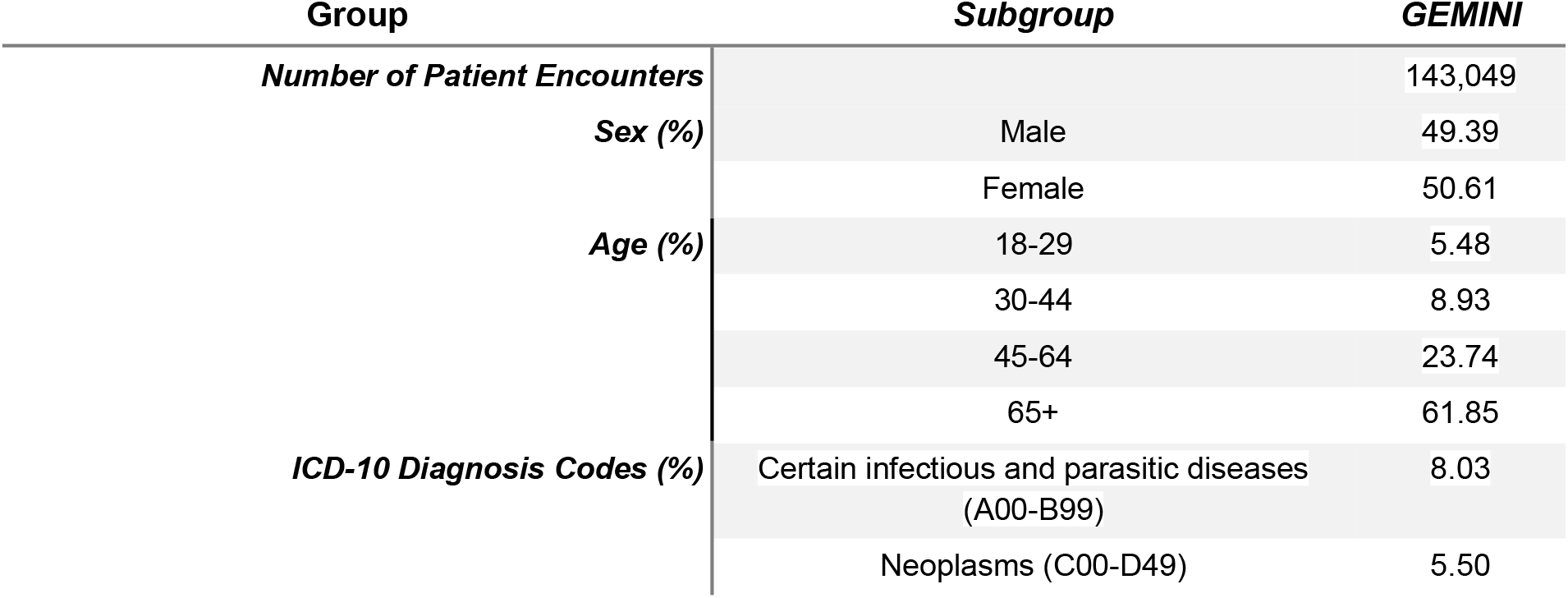
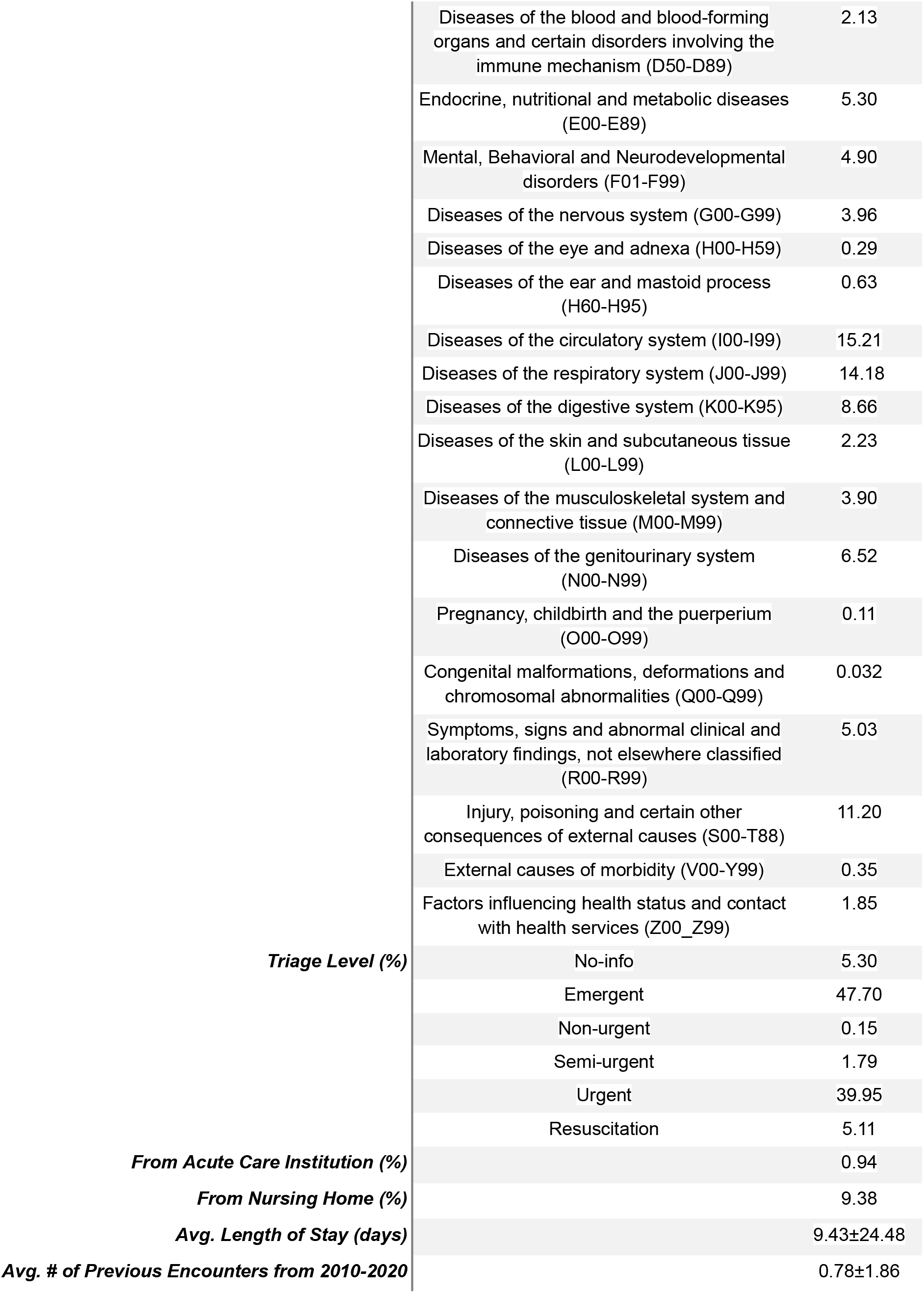
Patient Characteristics. Number of patient encounters, demographics, diagnoses, triage level, patient encounters from acute care institutions, patient encounters from nursing homes, average length of stay (LOS), and average number of previous encounters from 2010-2020, for GEMINI.

### Detection of harmful data shifts for the evaluation and monitoring of clinical AI systems

In the clinical setting, there are a myriad of factors that can contribute to a model drifting and making erroneous predictions, such as changes in behaviour, technology, population or policy ^18^. Using our monitoring and evaluation pipeline (**Figure 2**)^37^, we detected data shifts in a label-agnostic manner across increasing sample sizes for scenarios that we would expect to pose a threat to clinical AI systems during deployment, due to fundamental differences in patient populations. These scenarios consist of differences in demographics, hospital type, seasonality, time of day of hospital admission (i.e. day vs. night), time of week of hospital admission (i.e. weekday vs. weekend), and whether patients were admitted from an acute care institution or nursing home. Harmful data shifts were defined as those statistically significant between the source and target test data (p-value <0.05). We detected harmful data shifts and associated performance degradation in five scenarios: when transferring models trained on i) community hospitals to academic hospitals (**Figure 3AB**), ii) patients admitted during the day to patients admitted at night (**Figure 3AC**), iii) patients not admitted from nursing homes to patients admitted from nursing homes (**Figure 3AD**), iv) patients admitted from acute care institutions to patients admitted from non-acute care institutions (**Figure 3AD**) and v) patients admitted from non-acute care institutions to patients admitted from acute care institutions (**Figure 3AD**). Interestingly, we found many of these harmful data shifts were unidirectional, suggesting that there exists patterns among patient encounters in academic hospitals, during night admissions and among patients admitted from nursing homes that are not captured at community hospitals, during day admissions, and among patients admitted from outside of nursing homes, respectively. Harmful data shifts were not detected across seasons or sex (**Figure 3EF**). Although a harmful data shift was identified when evaluating on the 45-64 year-old age group, an associated decrease in AUROC did not occur (**Figure 3AG**).

**Figure 2.**
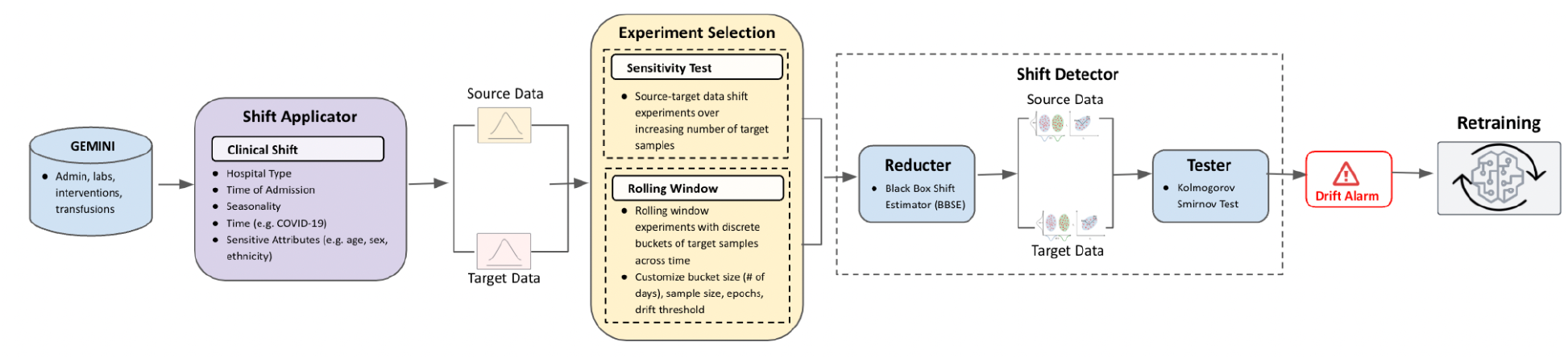
Monitoring and evaluation pipeline. An end-to-end pipeline where EHR data is first sent to the *Shift Applicator*, which outputs a source and target data based on the clinical shift of choice. The source and target data can then be leveraged by the *Shift Detector* to conduct a drift sensitivity test or a rolling window analysis–if drift is detected, retraining is triggered.

**Figure 3.**
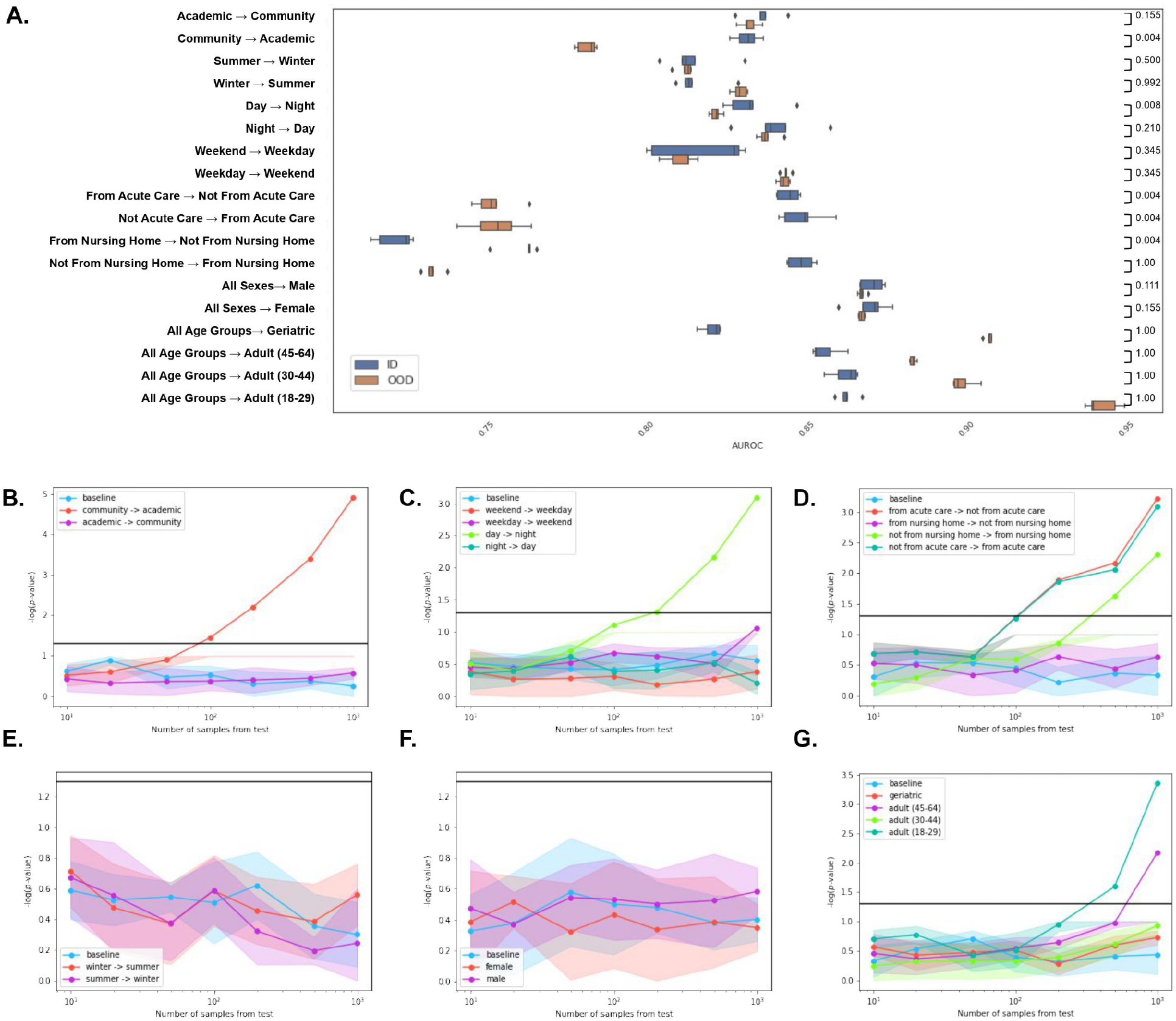
Detection of harmful data shifts. **(A)** Performance across in-distribution (ID) and out-of-distribution (OOD) data for demographics, hospital types, seasons, time of day of hospital admission, time of week of hospital admission, and whether the patient was admitted from an acute care institution or nursing home. P-values were calculated using a one-sided Mann-Whitney U test. Sensitivity of the data shift detection to an increasing number of test samples was evaluated for **(B)** hospital types **(C)** time of day or week of hospital admission **(D)** seasons **(E)** admission from acute care institutions or nursing homes **(F)** sex and **(G)** age. The horizontal black line represents a p-value threshold of 0.05.

These data shifts can arise for a variety of reasons, including differences in patient subpopulations, staffing, and/or resources that are not adequately represented in the training data ^38^. Assessing the fairness of the EWS for subgroups of diagnoses, sex and age across all the scenarios where harmful data shifts were identified, we found that there was decreased performance in numerous diagnostic subgroups between the source and target data (**Supplementary Figure 4**). The largest performance differences between patients from acute care and non-acute care institutions was for diseases of the nervous system (G00-G99), and musculoskeletal system and connective tissue (M00-M99). Between patients admitted during the day and night, the largest decrease in AUROC was seen in patients with neoplasms (C00-D49) and diseases of the musculoskeletal system and connective tissue (M00-M99) and genitourinary system (N00-N99). When transferring from community hospitals to academic hospitals, the largest performance decrease across diagnostic subgroups was for patients with neoplasms (C00-D49), which is also found at a much higher prevalence in academic hospitals (**Supplementary Table 3**). Alternatively, this may be due to differences in the 45-64 year-old age group, which significantly decreased in performance when models were transferred from community hospitals to academic hospitals (p=0.0079; **Supplementary Figure 4**).The hospital type shift could also be in part be driven by the increased number of individuals admitted from nursing homes in community hospitals compared to academic hospitals (**Supplementary Table 4**). This is also supported by our finding that models transferred from patients not admitted to nursing homes to patients admitted to nursing homes–which primarily consist of long-term care residents over the age of 85, result in harmful data shifts (**Figure 3AD**). It is also worth noting that the hospital type groupings coincide with differences in location which may also be a contributing factor of the data shift; more specifically, the academic hospitals are located in the central city while the community hospitals are located in residential suburbs. Interestingly, we found the inclusion of ICD-10 diagnosis codes as features decreased model deterioration due to data shifts (**Supplementary Table 5**).

### Preventing harmful data shifts during cross-site deployment

It is common practice that an ML model is developed at one institution and transferred to other institutions for external validation. During cross-site evaluation, we found that differences in hospital type result in harmful data shifts that deteriorate model performance (**Figure 3B**). In order to address this, we developed EWSs for i) each individual hospital, ii) the combination of community hospitals, iii) the combination of academic hospitals and iv) the combination of all hospitals. We then compared strategies leveraging a) *pre-training* where we used a model pre-trained on source data and evaluated it on out-of-distribution data from the target hospital, b) *transfer learning* where we fine-tuned the performance on the target hospital prior to evaluating the target data and c) *ablation* where we excluded data from a single hospital prior to evaluating the target data. For each model, we evaluated the performance for each individual hospital using a held out test set (**Figure 4A**). In general, cross-site training improved model performance; however, the use of all sites was never the optimal strategy suggesting that more data is not always helpful. We found training across all sites marginally improved model performance for academic hospitals but decreased performance for community hospitals (**Figure 4B**). Instead, using the model trained on both the community hospitals (Hospital 4 and 5) resulted in superior performance for community hospitals. Overall, fine-tuning the corresponding hospital type-specific model on the target hospital improved performance for all hospitals except Hospital 2. Interestingly, Hospital 2 is also the only hospital with a veteran’s wing, where patients receiving palliative care were less likely to experience in-hospital mortality and where the number of previous hospital visits was negatively correlated with risk of in-hospital mortality (**Supplementary Table 4**). In certain instances, the exclusion of a single hospital site improved model performance for another hospital. For Hospital 2, ablating Hospital 3 resulted in the best performing model (**Figure 4B**). It is worth noting, Hospital 2 and Hospital 3 also had the largest difference in the number of individuals with diseases due to factors influencing health status and contact with health services (17%), which is the diagnostic subgroup with the lowest performance (**Supplementary Figure 3**; **Supplementary Table 3**; **Figure 1E**). These two hospitals also had the largest difference in patients receiving palliative care between mortality status; Hospital 2 had a 2.7-fold decrease and Hospital 3 had a 6.3-fold increase in palliative care among patients who died in the hospital. The population demographic and socioeconomic status (SES) between the two hospitals are also very different; Hospital 3 is an inner city urban and Hospital 2 is a suburban hospital. It is important that clinical AI systems be proactively evaluated for these differences so they are considered when transferring models across sites.

**Figure 4.**
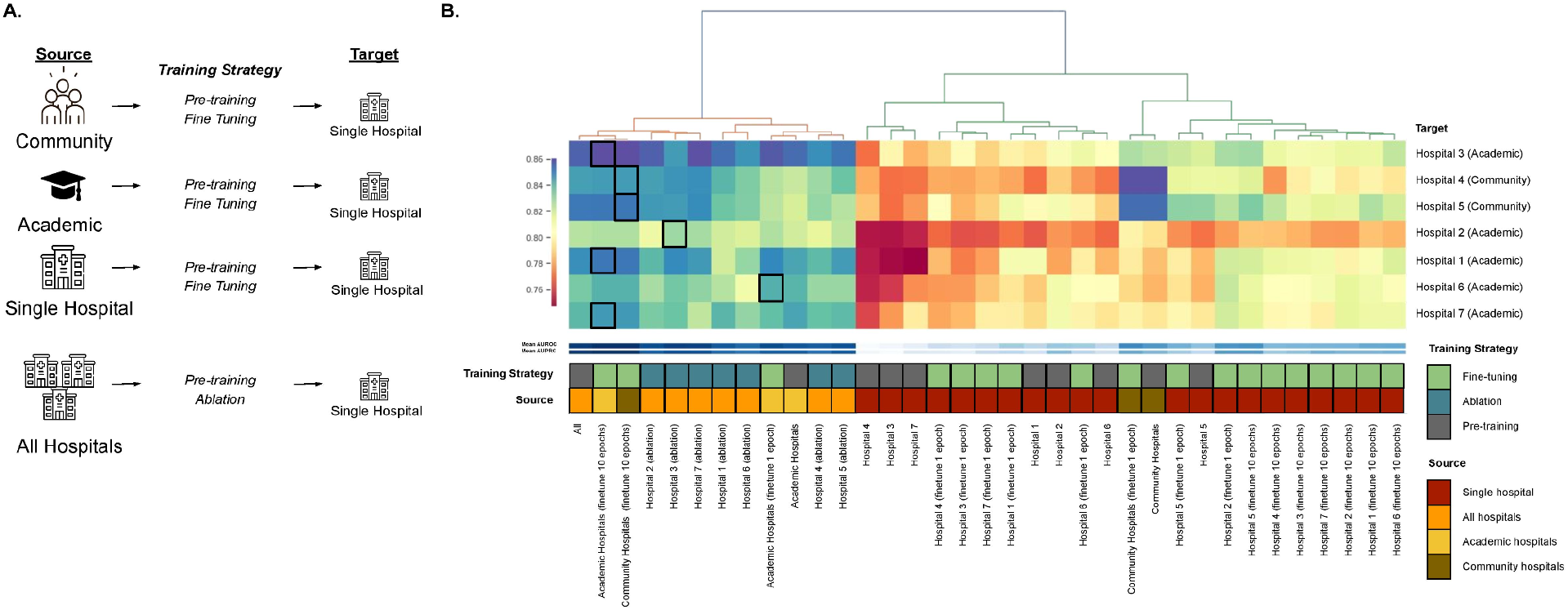
Optimal training strategies for cross-site deployment. **(A)** Pre-training, fine-tuning and ablation employed on single-site models, a cross-site model and hospital-type specific models (community, academic). **(B)** Heatmap of AUROC of the training strategies across each test hospital site. Highlighted in black is the best performing model for each hospital.

### Detecting and mitigating model deterioration due to temporal data shifts

Lastly, we conducted a simulated prospective evaluation of an EWS for mortality prediction using GIM data from 2010-2018. In a real-time deployment scenario, labels are not always readily available at the time of prediction. Moreover, for outcomes like mortality, the problem with relying on model performance is that the event rate is relatively rare, so it can take many months to accrue a sufficient sample size for detecting model performance changes. As a result, label-agnostic drift detection is critical for identifying model degradation and triggering retraining procedures. We monitored our EWS for temporal shifts using a 14-day rolling window from March 2019 to August 2020. In the presence of drift, we used continual learning strategies to update our model and mitigate model deterioration (**Figure 5A**). First, we compared *periodic retraining*–whereby the model is updated at regular, pre-defined intervals and *drift-triggered retraining*–whereby the model is updated when there is a significant data shift between the source data and target data. We found drift-triggered retraining resulted in better overall performance (**Supplementary Figure 5**). To identify the optimal approach for drift-triggered retraining, we tuned various parameters including the retraining window size, lookahead window, sample size, drift threshold and number of epochs (**Supplementary Figures 6-10**). The retraining window represents how much previous data we want to use for updating the model. We found a larger retraining window improved AUROC and AUPRC, however, as the retraining window increased upwards of 180 days, the performance decreased, suggesting that greater amounts of past data are not always beneficial for model updating (**Supplementary Figure 6**). Due to the lead time for acquiring labels, it is possible that at the time model updating is triggered, labels for the most recent patient encounters are not available. As a result, we evaluated increasing lookback window sizes to determine how far back the data used to update the model can be, without sacrificing performance. We found lookback windows of up to 60 days were able to maintain similar model performance (**Supplementary Figure 7**). Although, the lookback window will differ depending on the frequency of the prediction outcome and the progression of the drift over time (i.e. gradual versus sudden). Given that the model updating is triggered by drift detection, the sensitivity of the drift test will influence the overall performance. We found that the optimal drift threshold was a p-value of 0.01 (**Supplementary Figure 8)** and the optimal number of encounters for the drift test was 1000 (**Supplementary Figure 9**). However, it is important to recognize each prediction task and domain is unique, and as a result the generalizability of the optimal threshold will need to be evaluated on a case-by-case basis. We also found that increasing the number of epochs during model updating resulted in catastrophic forgetting whereby the model overfit and model performance decreased over time (**Supplementary Figure 10**). In addition, we compared updating whereby we only trained on encounters that were predicted correctly or positively; however, this was not as effective as using all the encounters (**Supplementary Figure 11**). Overall, the implementation of our drift-triggered continual updating strategy improved model performance over time and was more effective than maintaining a locked model during deployment (**Figure 5B**).

**Figure 5.**
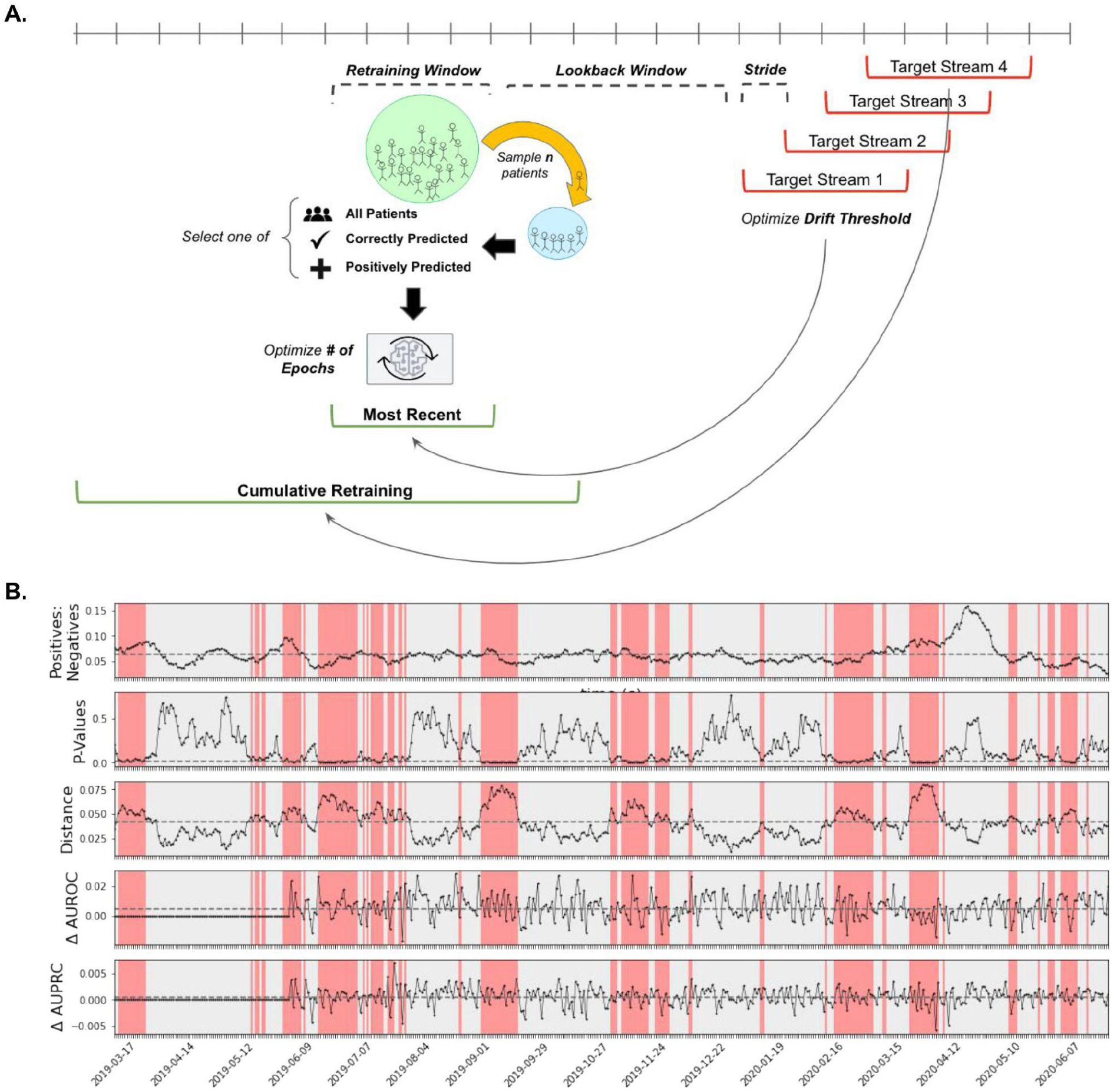
Prospective evaluation of EWS for mortality prediction. **A.** Model updating strategies and parameters explored whereby the target stream is evaluated on i) the most recent n days or ii) all the encounters seen to date, with the option to select training on only the encounters predicted positively or correctly. Each strategy can be optimized for drift threshold, number of samples, number of epochs, stride length, lookback window, and retraining window. **B.** We monitored the proportion of positive:negative outcomes, drift p-values, and drift distance metric from 03/2019 to 08/2020 using a 14-day rolling window.

In the event that drift is detected, model updating is triggered (red), for which we also monitored the change in AUROC and AUPRC between the retrained model and the baseline model, which implements no updating procedure.

## Discussion

Many widely implemented clinical AI systems ^26,39,40^, have demonstrated poor generalizability upon external validation, as a result of harmful data shifts. However, these biases are rarely accounted for in a proactive manner, and are typically identified following deployment, while relying on ground-truth labels ^18,41,42^. In this study, we built a dynamic EWS that adapts to the ever-changing healthcare environment. We used our EWS to predict the risk of mortality to enable the effective triaging of patients admitted to GIM and performed robust evaluations for bias and data shifts across diagnostic subgroups, demographics, hospital sites, based on the when and where a patient was admitted, and over time. We accurately detected harmful data shifts in clinical data without relying on ground-truth labels by leveraging black box shift detection and two sample testing^28^; this permitted the proactive evaluation of ML models in clinical settings where labels can be costly, resource-intensive, and delayed. In doing so, we found models trained on patients admitted during the day do not generalize well to patients admitted at night, emphasizing the importance of careful cohort selection for model development. We also found harmful data shifts attributed to whether or not a patient was admitted from an acute care institution or nursing home, suggesting these settings have distinct patient populations. Institutional differences are among the most common causes of data shifts due to underlying differences in patient demographics, disease incidence and data-collection workflows ^2,11^. We found models built on specific groups of hospitals such as community hospitals, undergo harmful data shifts when evaluated on academic hospitals and compared training strategies to mitigate model deterioration attributed to cross-site deployment. Lastly, we monitored data shifts over time and investigated key questions surrounding model updating like when to update a model, how much data to update on, and what data to use for the update. We found our drift-triggered continual updating strategy improved model performance and was more effective than maintaining a locked model during deployment.

However, it is unclear to what extent our findings will generalize, which is why it is critical to perform these experiments across several prediction tasks, patient populations and types of shifts. Likewise, many other sensitive attributes (e.g. socioeconomic status) and clinical scenarios (e.g. specialized hospitals) that merit evaluation remain. It is also imperative to characterize the extent to which other data modalities, like clinical notes, contribute to biases in clinical AI systems. There are a number of reasons these shifts could occur, including changes in the distribution of diagnoses, staffing, or resource allocation across patient populations. Identification of causal structures is a promising strategy to help explain the failures of fairness transfer across distribution shifts ^43^. Given the sensitivity of clinical data, it is also important that future drift detection and retraining strategies consider privacy-preserving methods to ensure institutional boundaries are respected and autonomy is maintained over patient data ^44–46^.

In this study, we developed a drift-triggered continual learning strategy to improve model performance over time. However, it is worth noting that continual learning is not without risks, including catastrophic forgetting and feedback loops ^47–49^. Unfortunately, our dataset is unable to fully capture these long-term trends, but as more data is accumulated it will become possible to understand the impact of these model updating strategies over extended periods of time. Another caveat is that the current regulatory state of continual learning systems does not clearly define how and what aspects of a clinical AI system are permitted to change following authorization^41^. There are also several other training and updating strategies we did not explore, which can be leveraged to improve model performance in the presence of data shifts, including domain generalization (DG)^50,51^, representation learning^13,52^, meta learning ^53,54^, and multi-task learning^55,56^. For instance, consideration of other relevant prediction tasks (e.g. LOS, ICU transfer)^55^ or patient populations^56^ for pre-training could improve model generalization. Similarly, DG methods have been used as an alternative to baseline empirical risk minimization (ERM), to mitigate data shift^57^. However, many DG methods have repeatedly only been shown to improve performance in the context of extreme synthetic shifts and demonstrate poor performance on real world EHR data ^58,59^.

Instead, alternative ERM approaches (i.e. those that use stratified training, balanced subpopulation sampling, or worst-case model selection) outperform DG methods and show promise in mitigating model bias ^50,51,60^. Unfortunately, many studies fail to consider strong and realistic ERM baselines.

Clinical AI systems are complex, and each will differ in its biases and optimal retraining and updating procedures. As such, we have developed a monitoring and evaluation pipeline as part of a broader ML operations (MLOps) framework for clinical AI systems^37^ to facilitate robust evaluation and monitoring prior to deployment. Too often clinical ML models are reported with high performance metrics, while being developed in isolation. It is important to ensure that models are designed with deployment in mind, to ensure the responsible deployment of clinical AI systems. We hope our work permits the robust evaluation and monitoring of clinical AI systems in an effort to bridge the gap between model development and deployment ^61–63^.

## Methods

### Cohort data

We conducted this study using de-identified Electronic Health Record (EHR) and hospital administrative data from 109,802 patients admitted to the general internal medicine (GIM) wards from 2010-2020 across 7 large hospitals in the Toronto, Canada-area. Participating hospitals included the following sites, which are academic and community hospitals affiliated with the University of Toronto: Mount Sinai Hospital, Toronto General Hospital, Toronto Western Hospital, Trillium Health Partners Mississauga, Trillium Health Partners Credit Valley, St. Michael’s Hospital, and Sunnybrook Health Sciences Centre. In order to avoid the disclosure of sensitive hospital information, in keeping with the conventions of the GEMINI data sharing network, we have anonymized hospital names in the presentation of results. To assess the data quality issues arising from data extraction and transfer procedure from hospital sites, a computational assessment of 7 data quality checks was performed followed by manual data validation. This rigorous internal validation process demonstrated 98-100% accuracy across key data types ^64^.

### Ethics approval

All patient data was collected and approved through GEMINI ^64,65^ under the oversight of the research ethics board (REB) at the Toronto Academic Health Science Network (REB reference number 15-087). The extension of the REB approval was issued by the Unity Health Toronto REB (reference number 15-087). A separate REB approval was obtained for Trillium Health Partners. All experiments were performed in accordance with institutional guidelines and regulations.

### Model features

The *base* model consisted of 91 features comprising laboratory tests, blood transfusions, imaging reports and administrative features (**Supplementary Table 1**). The *base+CM* model consisted of the 91 features used in our base model, in addition to 18 comorbidities derived using ICD-10 codes (**Supplementary Table 1**). The *base+DxC* model consisted of the 91 features used in our base model, in addition to the 22 groupings of ICD-10 diagnosis codes (**Supplementary Table 1**). The input features used for time-series modelling were aggregated by taking the mean for 24-hour timesteps, over 144 hours.

### All-cause in-hospital mortality decompensation prediction

Our goal was to predict whether the patient’s health will rapidly deteriorate ^55^. Each instance of this task is a binary classification instance and predictions are made every 24 hours for the risk of in-hospital mortality within the next two weeks starting 24 hours after admission using the target replication approach (**Supplementary Figure 1**)^66^. In addition to longitudinal clinical measures, demographics are included as static variables at every time step for the prediction task. Labels were encoded as 1 if a patient died within the next 2 weeks, 0 if they were alive within the next 2 weeks and -1 if they were discharged. Missing values were imputed using forward filling followed by backward filling. We conducted a simulated deployment using a 8:2 random split of data from 2010-2018 for training and validation, and 2019-2020 data for testing.

### Clinical data shift experiments

We used prior knowledge to devise data splits that reflect real-life scenarios that may result in harmful data shifts and model degradation of clinical AI systems. For each experiment we trained our model on the source data, and evaluated on in-distribution (ID) source data as the *baseline* and out-of-distribution (OOD) target data as the *shift experiment*. More specifically, the source and target data were each split into 6:2:2 for training, validation and testing, respectively. For both the *baseline* and *shift experiment*, a model was trained on 60% of the source data. For the *baseline*, the model was fit and tuned on the ID validation data (20%) and evaluated on the ID test data (20%). For the *shift experiment*, the model was fit and tuned on the OOD validation data (20%) and evaluated on the OOD test data (20%). Each split is random and there were no overlapping patient encounters between the training, validation and test sets. All reported performance results are from the test set.

The source and target data for the scenarios are as follows:

**Winter** - *Source (ID)*: Patients admitted in the winter (Nov-Feb). *Target (OOD)*: Patients admitted in the winter (June-Aug).

**Summer** - *Source (ID):* Patients admitted in the summer (June-Aug) *Target (OOD):* Patients admitted in the winter (Nov-Feb).

**Community hospitals** - *Source (ID):* Academic hospitals (Hospital 1, Hospital 2, Hospital 3, Hospital 6, Hospital 7). *Target (OOD):* Community hospitals (Hospital 4, Hospital 5).

**Academic hospitals** - *Source (ID):* Community hospitals (Hospital 4, Hospital 5).*Target (OOD)*: Academic hospitals (Hospital 1, Hospital 2, Hospital 3, Hospital 6, Hospital 7).

**Day admission** - *Source (ID):* Patients admitted during the day (7:30-19:30). *Target (OOD)*: Patients admitted during the night (0:00-7:30,19:30:23:59).

**Night admission** - *Source (ID):* Patients admitted during the night (0:00-7:30,19:30:23:59). *Target (OOD)*: Patients admitted during the day (7:30-19:30).

**Weekend admission** - *Source (ID):* Patients admitted on the weekend (i.e. Saturday and Sunday). *Target (OOD)*: Patients admitted on a weekday (i.e. Monday to Friday).

**Weekday admission** -*Source (ID):* Patients admitted on a weekday (i.e. Monday to Friday). *Target (OOD)*: Patients admitted on the weekend (i.e. Saturday and Sunday).

**Admitted from nursing home**- *Source (ID):* Patients admitted from nursing homes. *Target (OOD)*: Patients not admitted from nursing homes.

**Not admitted from nursing home** - *Source (ID):* Patients not admitted from nursing homes. *Target (OOD)*: Patients admitted from nursing homes.

**Admitted from acute care institution** - *Source (ID):* Patients admitted from acute care institutions. *Target (OOD)*: Patients not admitted from acute care institutions.

**Not admitted from acute care institution** -*Source (ID):* Patients not admitted from acute care institutions. *Target (OOD)*: Patients admitted from acute care institutions.

**Sex** - *Source (ID):* Patients of all sexes. *Target (OOD)*: Patients that are i) males ii) females.

**Age** - *Source (ID):* Patients of all ages. *Target (OOD)*: Patients that are i) 18-29 years ii) 30-44 years iii) 45-64 years iv) 65+ years.

### Model training and evaluation

The training, validation and test data were normalized independently by subtracting the mean and scaling to unit variance. A recurrent neural network (RNN), gated recurrent unit (GRU), and long short-term memory (LSTM) ^66^ with 2 hidden layers, 64 hidden cells and a dropout rate of 0.2 were implemented using PyTorch^67^. The time-series models were optimized for binary cross entropy with logits loss using Adagrad^68^, a step size of 128, gamma of 0.5, learning rate of 3.0 × 10^−2^, weight decay of 1.0 × 10^−6^ and batch size of 64. To account for the class imbalance, we reweighted our loss function by the fraction of controls/cases in the training data. Each model was trained over 128 epochs with early stopping using a patience of 3 and delta of 0. We used a sigmoid activation function to obtain prediction probabilities. We generated standard errors by making a random choice of weight initializations and performed 5 repetitions for each dataset split. We used the area under the receiver operator characteristic curve (AUROC) and area under the precision recall curve (AUPRC) to evaluate performance, and sensitivity and positive predictive value (PPV) to evaluate clinical utility. The LSTM had the highest performance and thus was used for downstream monitoring and evaluation (**Supplementary Table 2**). Model parameters (e.g. number of cells, number of layers) were kept fixed throughout the experiments.

### Monitoring and evaluation pipeline

We detected distributional shifts between source and target data using our monitoring and evaluation pipeline (**Figure 2**) which consists of:

1. **Shift application**: EHR data is sent to the *Shift Applicator*, which outputs a source and target dataset based on the clinical data shift experiment of choice (e.g. hospital type, seasons, etc.).
2. **Dimensionality reduction**: Dimensionality reduction is performed using the *Shift Reductor* to obtain a latent representation of the source and target data. This was done using the softmax outputs of a LSTM neural network label classifier trained on source data (Black Box Shift Detector; BBSD) ^69^. The architecture and training of the BBSD is described above as the base model. For each data shift experiment, an LSTM model trained on the source data (as described above) was used as the BBSD.
3. **Statistical testing:** Univariate two-sample testing was performed with a Kolmogorov-Smirnov Test using the *Shift Tester*, in order to identify if a harmful data shift has occurred between the latent representation of the source and target data^28^.
4. **Sensitivity test:** A drift sensitivity test was conducted by performing step (2) and (3) to detect data shifts for n = {10, 20, 50, 100, 250, 500, 1000} patients from the target data.
5. **Rolling window analysis:** A 14-day rolling window was used to assess model stability over time by sampling 1000 patients and performing step (2) and (3) to test for drift every day. The drift detector was updated every day with the last 25000 patients.

### Training strategies for cross-site deployment

To evaluate the optimal training strategy for each hospital, we trained models using i) each individual hospital, ii) the combination of hospitals for each hospital type (i.e. academic, community) and ii) all hospitals. For each LSTM model, the data for the corresponding hospital(s) were partitioned into random splits of 6:2:2 for training, validation and testing. We compared strategies including i) *pre-training* where we used a model pre-trained on source data and evaluated it on test data from the target hospital, ii) *fine-tuning* where the single-site and hospital-type specific models were fine-tuned on the validation set for the target hospital by freezing all layers except the final layer and using 1 epoch or 10 epochs, prior to evaluation on test data from the target hospital, and iii) *ablation* of a single hospital from the cross-site model, prior to evaluation on test data from the target hospital. The performance of each strategy was evaluated on test sets consisting of 20% of data, from each of the 7 individual hospital sites.

### Prospective evaluation and continual learning

We evaluated performance on test data every day (stride=1) from 03/2019 to 08/2020 using a 14-day rolling window. In order to mitigate model drift due to temporal data shifts, we compared a baseline where the model was kept locked and no changes or updates were made, to the following continual learning strategies:

**Periodic updating** - The model is updated at regular time intervals of n = {7, 14, 30, 60} days (**Supplementary Figure 5**).

**Most-recent updating**- When drift is detected, the model is updated using the most recent n number of days where n = {7, 14, 30, 60, 120, 180, 270} days (**Supplementary Figure 6**).

**Cumulative updating** - When drift is detected, the model is updated using all the patient encounters seen to-date.

Model updating methods were optimized for the retraining window size (n = {7, 14, 30, 60, 120, 180, 270} days), lookback window (**Supplementary Figure 7**; n = {0, 30, 60, 120} days), drift p-value threshold (**Supplementary Figure 8**; p = {0.1, 0.05, 0.01, 0.001}), sample size used for the drift test (**Supplementary Figure 9**; n = {50, 100, 250, 500, 1000, 1500}), and number of epochs (**Supplementary Figure 10**; n = {1, 5, 10}). We also compared sampling strategies where we used i) all the encounters in the retraining window ii) only the correctly predicted encounters in the retraining window and ii) only the positively predicted encounters in the retraining window.

## Data Availability

Access to GEMINI data for research purposes can be requested by a submission of a Project Proposal Form for review by the GEMINI Projects and Publications Committee. Further information can be found at https://www.geminimedicine.ca/access-data.

## Code Availability

The code used to perform the experiments in this study can be found at https://github.com/vsubasri/GEMINI-data-shift. We have also provided our monitoring pipeline as part of a broader MLOps framework, to facilitate the research and deployment of ML models in the clinical setting ^37^.

## Data Availability

All data produced in the present study are available to authors with access to GEMINI, upon reasonable request.

## Acknowledgements

Acknowledgements

This work is made possible due to the data obtained from the General Medicine Inpatient Initiative (GEMINI), and we acknowledge the GEMINI team for their support. We also acknowledge HPC4Health, for enabling high performance computing environments which were involved in the development of this work. Finally, we acknowledge support from the Vector Institute and its vibrant community working at the intersection of health and machine learning. V.S. is supported by Ontario Graduate Scholarship and a Vector Institute grant. A.V. is supported by the Temerty Professorship in Artificial Intelligence Research and Education in Medicine at the University of Toronto. D.M. is supported by the CIBC Children’s Foundation Chair in Child Health Research. A.G. is supported by the Varma Family Chair and CIFAR AI Chair.

