## Supplemental Materials for "Diagnosing and remediating harmful data shifts for the responsible deployment of clinical AI models"

### Supplementary Materials

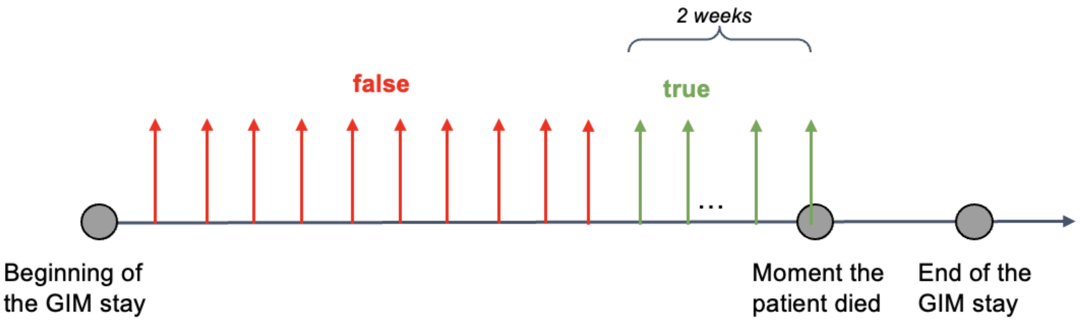


**Supplementary Figure 1. Mortality decompensation prediction in general internal medicine (GIM).** Model predicts whether patients' health deteriorates and results in hospital mortality within the next 2 weeks, every 24 hours of a GIM stay.

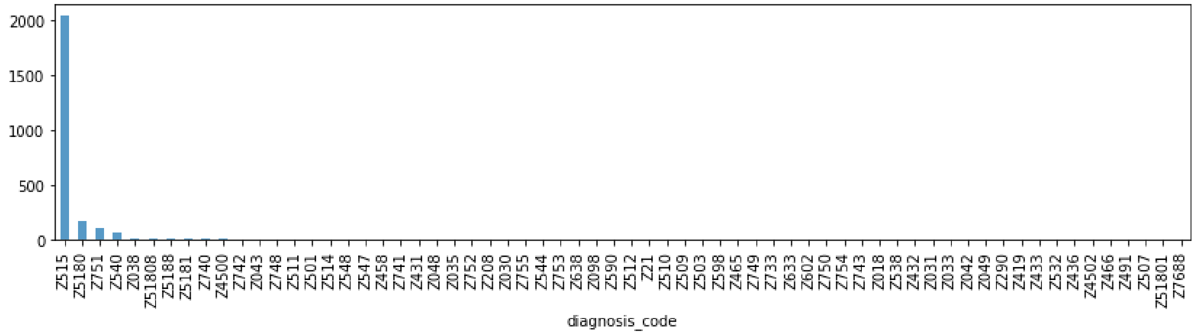


**Supplementary Figure 2.** Distribution of diagnosis codes across factors influencing health status and contact with health services (Z00-Z99).

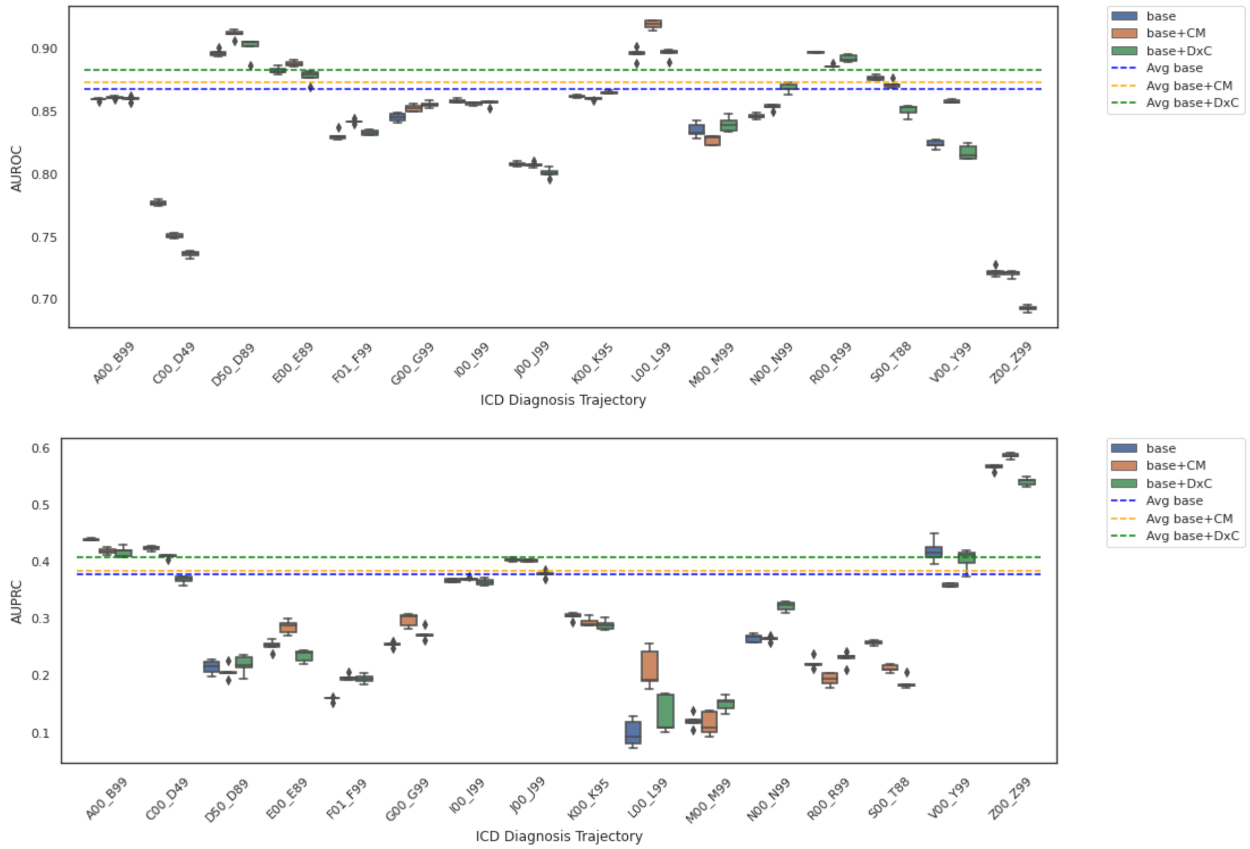


**Supplementary Figure 3.** Performance of model using no prior information (base), comorbidities (base+CM) and ICD-10 diagnosis codes (base+DxC) across diagnosis codes measured using AUROC and AUPRC. The dotted line represents the average performance for each respective model, across all groups of ICD-10 diagnosis codes.

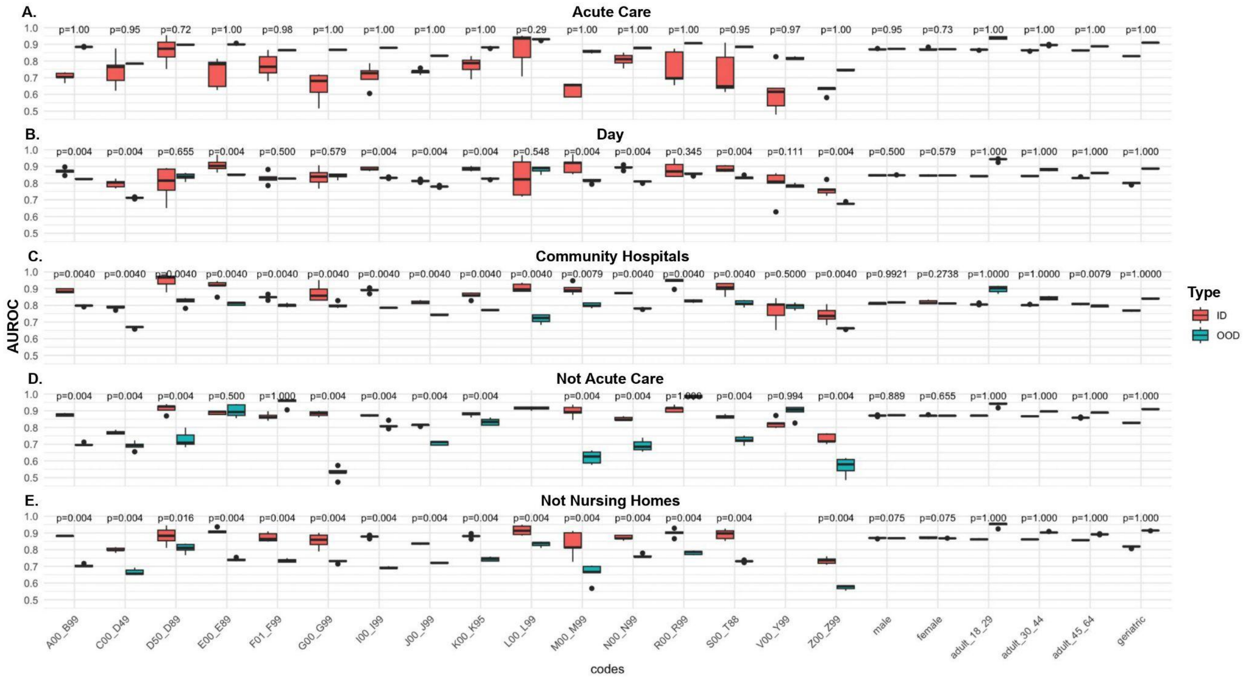

**Supplementary Figure 4.** AUROC for in-distribution (ID) and out-of-distribution (OOD) data, across ICD-10 diagnosis codes, age and sex, for scenarios where harmful data shifts were detected: **(A)** model trained on patients admitted from acute care institutions **(B)** model trained on patients admitted during the day **(C)** model trained on patients admitted from community hospitals **(D)** model trained on patients admitted not from acute care institutions **(E)** model trained on patients not admitted from nursing homes. P-values were calculated using a one-sided Mann-Whitney U test.

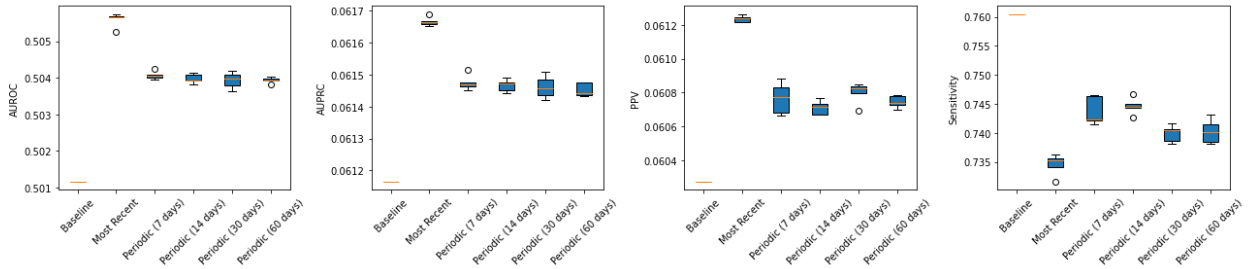

**Supplementary Figure 5.** Comparison of AUROC, AUPRC, PPV, and sensitivity when updating periodically every n = 7, 14, 30, and 60 days.

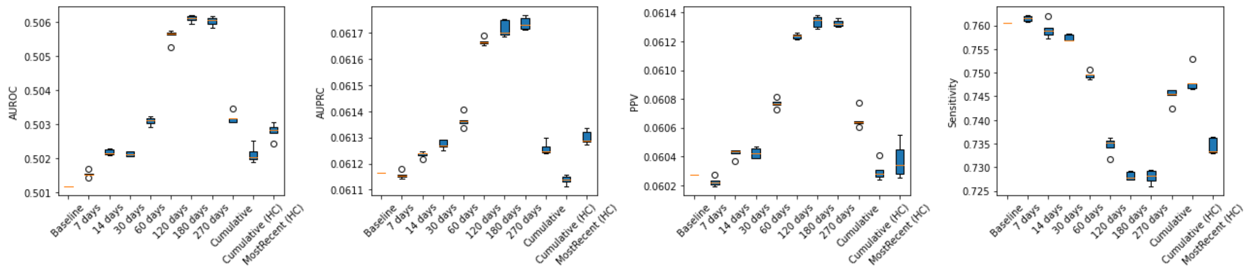

**Supplementary Figure 6.** Comparison of AUROC, AUPRC, PPV, and sensitivity across strategies retraining using a dynamic window of the most recent encounters (n = 7, 14, 30, 60, 120, 180, 270 days) and cumulatively.

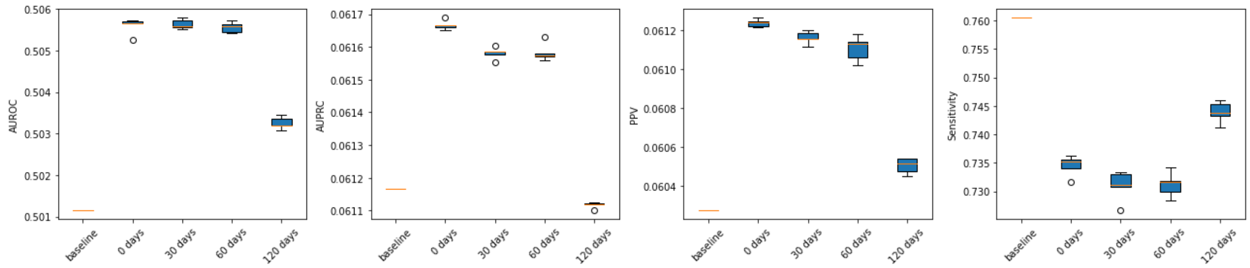


**Supplementary Figure 7.** Comparison of AUROC, AUPRC, PPV, and sensitivity across increasing lookback windows (n = 0, 30, 60, 120 days).

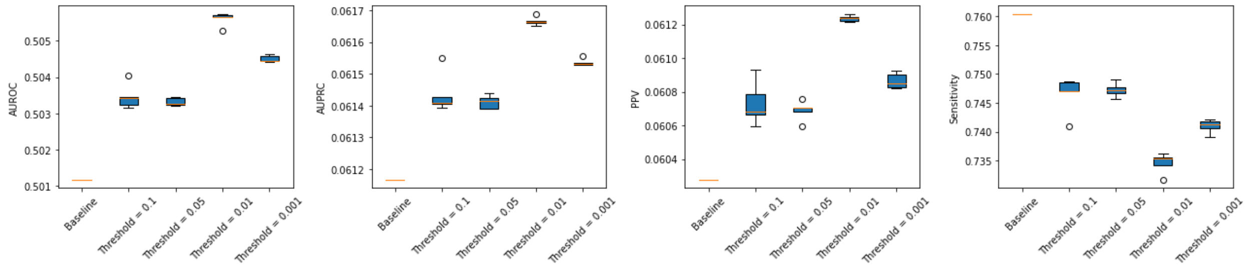


**Supplementary Figure 8.** Comparison of AUROC, AUPRC, PPV, and sensitivity across varying drift p-value thresholds for retraining (p = 0.1, 0.05, 0.01, 0.001).

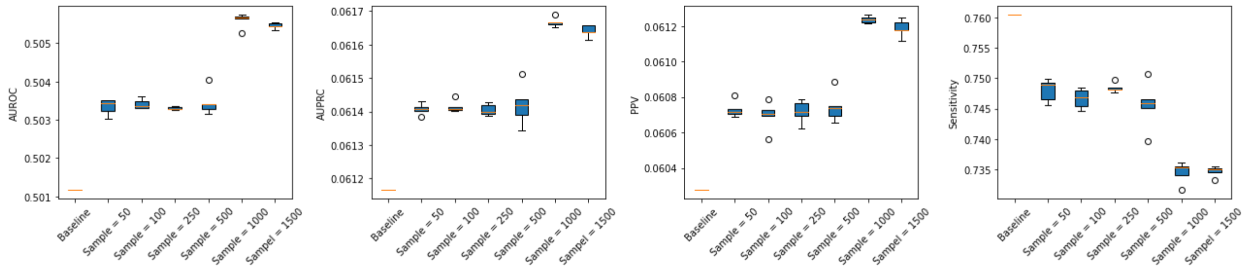


**Supplementary Figure 9.** Comparison of AUROC, AUPRC, PPV, and sensitivity across varying sample sizes for drift tests (n = 50, 100, 250, 500, 1000, 1500).

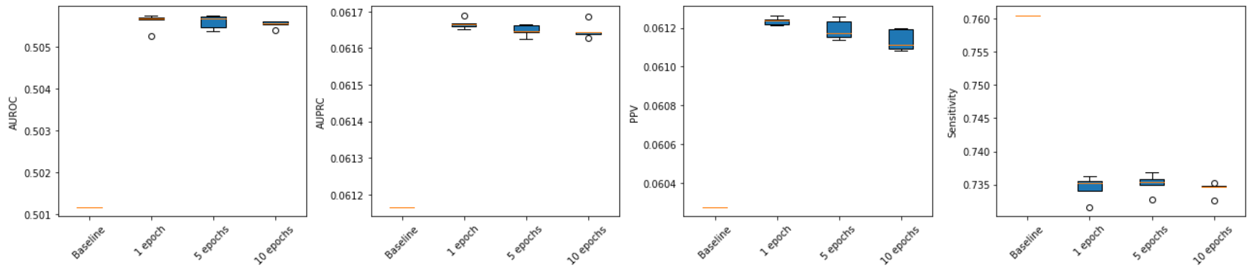


**Supplementary Figure 10.** Comparison of AUROC, AUPRC, PPV, and sensitivity across varying epochs for retraining (n = 1, 5, 10).

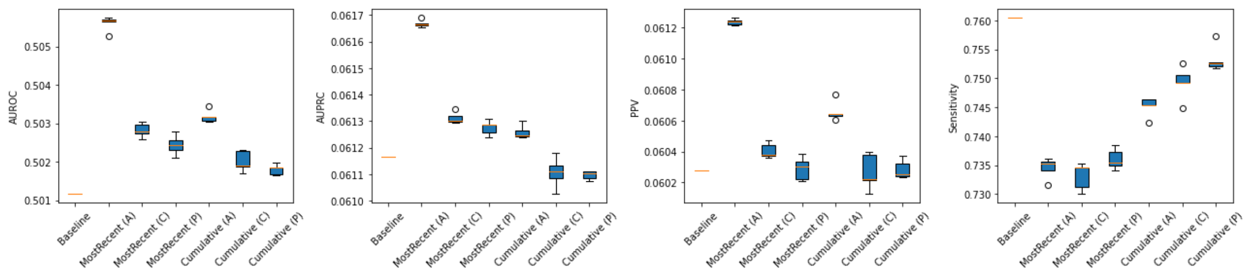


**Supplementary Figure 11.** Comparison of AUROC, AUPRC, PPV, and sensitivity when updating using the most recent and cumulative retraining strategy with all encounters (A), correctly predicted encounters (C) or positively predicted encounters (P).

| Feature Type | # of Features | Features |
|---|---|---|
| Administrative | 16 | sex, age, prev_encounter_count, triage_level_emergent, triage_level_no_info, triage_level_non-urgent, triage_level_resuscitation, triage_level_semi-urgent, triage_level_urgent, readmission_new_to_acute, readmission_nota, readmission_planned_from_acute, readmission_unplanned_7_day_acute, readmission_unplanned_7_day_day_surg, readmission_unplanned_8_to_28_day_acute, from_nursing_home_mapped, from_acute_care_institution_mapped |
| Interventions | 6 | unmapped_intervention, inv_mech_vent_mapped, endoscopy_mapped, dialysis_mapped, surgery_mapped, interventional |
| Labs | 55 | albumin, alp, alt, aptt, arterial paco2, arterial pao2, arterial ph, ast, bicarbonate, bilirubin, blood urea nitrogen, calcium, calcium ionized, creatinine, crp, d-dimer, esr, ferritin, fibrinogen, glucose fasting, glucose point of care, glucose random, hba1c, hematocrit, hemoglobin, high sensitivity troponin, influenza, inr, ketone, lactate arterial, lactate venous, ldh, lipase, lymphocyte, mean cell volume, neutrophils, other, platelet count, potassium, pt, serum alcohol, serum osmolality, sodium, troponin, tsh, urinalysis, urine osmolality, urine sodium, urine specific gravity, venous pco2, venous ph, vitamin b12, vitamin d, white blood cell count |
| Imaging Reports | 5 | ct, mri, x-ray, echo, ultrasound |
| Blood Transfusions | 2 | rbc, non-rbc |
| Comorbidities<br><br>*(Only used in Base+CM)* | 18 | Kidney disease: N18, N19<br>Ischemic heart disease: I20-I52<br>Cerebrovascular disease: I60-69<br>Hypertension: I10-I15<br>Diabetes: E10-E13<br>Hyperlipidemia: E78<br>Hypertension: I10<br>Congestive heart failure: I50<br>Cancer: C00-D49<br>Dyspnea: R06<br>COPD: J44<br>Asthma: J45<br>Pulmonary embolism: I26<br>Connective tissue disease: I30-I36<br>Inflammatory bowel disease: K50, K51,<br>Osteoarthritis: M15-M19 |

| | | |
|---|---|---|
| | | Rheumatoid arthritis: M05-M14<br>HIV: B20-B24 |
| ICD-10 Diagnosis Codes<br><br>*(Only used in Base+DxC)* | 22 | Certain infectious and parasitic diseases: A00-B99<br>Neoplasms: C00-D49<br>Diseases of the blood and blood-forming organs and certain disorders involving the immune mechanism: D50-D89<br>Endocrine, nutritional and metabolic diseases: E00-E89<br>Mental, Behavioral and Neurodevelopmental disorders: F01-F99<br>Diseases of the nervous system: G00-G99<br>Diseases of the eye and adnexa: H00-H59<br>Diseases of the ear and mastoid process: H60-H95<br>Diseases of the circulatory system: I00-I99<br>Diseases of the respiratory system: J00-J99<br>Diseases of the digestive system: K00-K95<br>Diseases of the skin and subcutaneous tissue: L00-L99<br>Diseases of the musculoskeletal system and connective tissue: M00-M99<br>Diseases of the genitourinary system: N00-N99<br>Pregnancy, childbirth and the puerperium: O00-O99<br>Certain conditions originating in the perinatal period: P00-P96<br>Congenital malformations, deformations and chromosomal abnormalities: Q00-Q99<br>Symptoms, signs and abnormal clinical and laboratory findings, not elsewhere classified: R00-R99<br>Injury, poisoning and certain other consequences of external causes: S00-T88<br>External causes of morbidity: V00-Y99<br>COVID19: U07-U08<br>Factors influencing health status and contact with health services: Z00-Z99 |

**Supplementary Table 1**. EHR features used for mortality decompensation prediction.

| Model | AUROC | AUPRC | Sensitivity | PPV |
|---|---|---|---|---|
| Recurrent neural network (RNN) | 0.835 ± 0.0005 | 0.328 ± 0.0026 | 0.887 ± 0.0030 | 0.162 ± 0.0016 |
| Gated recurrent unit (GRU) | 0.838 ± 0.0013 | 0.344 ± 0.0030 | 0.865 ± 0.0067 | 0.177 ± 0.0008 |
| Long short-term memory (LSTM) | 0.846 ± 0.0009 | 0.349 ± 0.0022 | 0.887 ± 0.0045 | 0.173 ± 0.0013 |

**Supplementary Table 2**. AUROC, AUPRC, sensitivity, and PPV of the test set, for time series models including RNN, GRU and LSTM models.

| Mortality | False | | | | | | | True | | | | | | |
|---|---|---|---|---|---|---|---|---|---|---|---|---|---|---|
| Hospital | 1(A) | 2(A) | 3(A) | 4(C) | 5(C) | 6(A) | 7(A) | 1(A) | 2(A) | 3(A) | 4(C) | 5(C) | 6(A) | 7(A) |
| A00_B99 | 0.086 | 0.065 | 0.065 | 0.078 | 0.082 | 0.083 | 0.072 | 0.152 | 0.115 | 0.099 | 0.135 | 0.112 | 0.119 | 0.129 |
| C00_D49 | 0.065 | 0.059 | 0.039 | 0.036 | 0.029 | 0.066 | 0.035 | 0.177 | 0.125 | 0.111 | 0.080 | 0.073 | 0.190 | 0.102 |
| D50_D89 | 0.032 | 0.018 | 0.019 | 0.015 | 0.016 | 0.048 | 0.018 | 0.011 | 0.004 | 0.003 | 0.004 | 0.003 | 0.015 | 0.007 |
| E00_E89 | 0.063 | 0.050 | 0.063 | 0.058 | 0.051 | 0.055 | 0.066 | 0.017 | 0.025 | 0.019 | 0.028 | 0.018 | 0.020 | 0.023 |
| F01_F99 | 0.047 | 0.047 | 0.053 | 0.052 | 0.072 | 0.026 | 0.057 | 0.020 | 0.032 | 0.031 | 0.030 | 0.048 | 0.013 | 0.023 |
| G00_G99 | 0.038 | 0.051 | 0.038 | 0.065 | 0.039 | 0.026 | 0.028 | 0.011 | 0.031 | 0.028 | 0.028 | 0.031 | 0.008 | 0.012 |
| H00_H59 | 0.004 | 0.006 | 0.003 | 0.005 | 0.001 | 0.001 | 0.002 | NA | NA | NA | NA | NA | NA | NA |
| H60_H95 | 0.006 | 0.006 | 0.004 | 0.012 | 0.009 | 0.006 | 0.006 | NA | NA | NA | NA | NA | NA | NA |
| I00_I99 | 0.101 | 0.147 | 0.143 | 0.138 | 0.219 | 0.138 | 0.131 | 0.103 | 0.211 | 0.165 | 0.175 | 0.249 | 0.119 | 0.158 |
| J00_J99 | 0.127 | 0.132 | 0.144 | 0.130 | 0.106 | 0.137 | 0.163 | 0.217 | 0.238 | 0.175 | 0.256 | 0.229 | 0.197 | 0.258 |
| K00_K95 | 0.108 | 0.103 | 0.118 | 0.059 | 0.046 | 0.106 | 0.094 | 0.051 | 0.081 | 0.079 | 0.048 | 0.039 | 0.085 | 0.064 |

| | | | | | | | | | | | | | | |
|---|---|---|---|---|---|---|---|---|---|---|---|---|---|---|
| **L00_L99** | 0.028 | 0.026 | 0.029 | 0.024 | 0.021 | 0.022 | 0.022 | 0.003 | 0.005 | 0.003 | 0.004 | 0.006 | 0.004 | 0.001 |
| **M00_M99** | 0.044 | 0.041 | 0.049 | 0.053 | 0.036 | 0.036 | 0.043 | 0.009 | 0.008 | 0.011 | 0.011 | 0.007 | 0.006 | 0.006 |
| **N00_N99** | 0.059 | 0.066 | 0.062 | 0.076 | 0.085 | 0.057 | 0.063 | 0.025 | 0.042 | 0.030 | 0.054 | 0.057 | 0.035 | 0.039 |
| **O00_O99** | 0.002 | 0.001 | 0.001 | 0.003 | 0.001 | 0.000 | 0.001 | NA | NA | NA | NA | NA | NA | NA |
| **Q00_Q99** | NA | NA | NA | NA | NA | NA | NA | NA | NA | NA | NA | NA | NA | NA |
| **R00_R99** | 0.094 | 0.134 | 0.120 | 0.113 | 0.116 | 0.142 | 0.129 | 0.019 | 0.030 | 0.028 | 0.011 | 0.030 | 0.036 | 0.035 |
| **S00_T88** | 0.074 | 0.038 | 0.041 | 0.072 | 0.061 | 0.036 | 0.059 | 0.027 | 0.023 | 0.016 | 0.033 | 0.031 | 0.016 | 0.018 |
| **V00_Y99** | 0.002 | 0.002 | 0.004 | 0.005 | 0.003 | 0.003 | 0.004 | 0.004 | 0.003 | 0.004 | 0.013 | 0.011 | 0.005 | 0.009 |
| **Z00_Z99** | 0.020 | 0.008 | 0.004 | 0.006 | 0.005 | 0.011 | 0.008 | 0.156 | 0.027 | 0.197 | 0.089 | 0.054 | 0.131 | 0.117 |

**Supplementary Table 3.** Proportion of patient encounters across ICD-10 diagnosis codes, hospital and mortality status. Values for groupings of diagnosis codes with < 5 patient encounters have been omitted due to privacy preserving practices required by GEMINI. A = academic hospital, C = community hospital.

| **Mortality** | **False** | | | | | | | **True** | | | | | | |
|---|---|---|---|---|---|---|---|---|---|---|---|---|---|---|
| **Hospital** | **1(A)** | **2(A)** | **3(A)** | **4(C)** | **5(C)** | **6(A)** | **7(A)** | **1(A)** | **2(A)** | **3(A)** | **4(C)** | **5(C)** | **6(A)** | **7(A)** |
| **# of Encounters** | 16620 | 27394 | 16100 | 15096 | 22691 | 16238 | 14072 | 2405 | 2306 | 1589 | 1415 | 3524 | 1791 | 1808 |
| **LOS (days)** | 8.47 | 8.44 | 7.90 | 8.73 | 10.05 | 7.55 | 9.03 | 14.59 | 17.19 | 17.62 | 19.02 | 17.72 | 12.80 | 14.12 |
| **# of Previous Encounters from 2010-2020** | 0.72 | 0.60 | 1.19 | 0.68 | 0.60 | 0.82 | 0.90 | 1.28 | 1.12 | 1.53 | 1.36 | 1.16 | 1.31 | 1.63 |
| **From Acute Care (%)** | 2 | 0 | 1 | 1 | 0 | 1 | 1 | 7 | 1 | 1 | 3 | 0 | 1 | 0 |
| **From Nursing Home (%)** | 6 | 8 | 4 | 11 | 12 | 4 | 10 | 13 | 18 | 14 | 31 | 30 | 8 | 22 |
| **Palliative Care (n)** | 260 | 151 | 50 | 52 | 39 | 160 | 80 | 373 | 56 | 313 | 123 | 191 | 235 | 210 |

**Supplementary Table 4. Patient admission and stay characteristics by hospital and mortality status.** Number of patient encounters, average length of stay (LOS), average number of previous encounters from 2010-2020, percentage of patient encounters from acute care institutions, percentage of patient encounters from nursing homes, and number of patients receiving palliative care, across hospitals and mortality status. A = academic hospital, C = community hospital.

| | | | **Base** | **Base + Comorbidities** | **Base + ICD-10 Diagnosis Codes** |
|---|---|---|---|---|---|
| **Source** | **Target** | **Metric** | **Target – Source (%)** | **Target – Source (%)** | **Target – Source (%)** |
| **Community Hospitals** | **Academic Hospitals** | AUROC | -4.0 | -2.6 | -0.9 |
| | | AUPRC | -6.8 | -5.0 | -3.2 |
| **Academic Hospitals** | **Community Hospitals** | AUROC | -1.1 | -1.8 | 0.0 |
| | | AUPRC | -3.2 | -4.8 | 0.0 |
| **Excl. Winter** | **Seasonal Winter** | AUROC | +0.7 | -0.3 | -1.2 |
| | | AUPRC | -0.6 | +0.9 | -0.1 |
| **Excl.** | **Seasonal** | AUROC | +0.8 | +1.2 | +0.3 |

| | | | | | |
|---|---|---|---|---|---|
| **Summer** | **Summer** | AUPRC | +1.0 | +0.9 | -0.3 |
| **Night Admission** | **Day Admission** | AUROC | +1.6 | -0.2 | +0.2 |
| | | AUPRC | +3.0 | +0.9 | +2.1 |
| **Day Admission** | **Night Admission** | AUROC | -1.9 | -0.7 | -0.8 |
| | | AUPRC | -5.6 | -3.2 | -3.7 |

**Supplementary Table 5**. Performance of models on in-distribution (source) and out-of-distribution (target) data without prior information (base), with comorbidities (base+CM) and with diagnosis codes (base+DxC) as features.
